# Considerations for Equitable Distribution of Digital Healthcare for People Who Use Drugs

**DOI:** 10.1101/2024.08.19.24312251

**Authors:** Zoi Papalamprakopoulou, Sotirios Roussos, Elisavet Ntagianta, Vasiliki Triantafyllou, George Kalamitsis, Arpan Dharia, Vana Sypsa, Angelos Hatzakis, Andrew H. Talal

## Abstract

**Background:** Telehealth holds the potential to expand healthcare access for people who use drugs (PWUD). However, approaches to increase PWUDs’ access to digital healthcare are not well-understood. We studied digital healthcare accessibility among PWUD.

**Methods:** We employed respondent-driven sampling to recruit 162 PWUD in Athens, Greece to collect data via a structured questionnaire. Participants were aged at least 18 years and had an injection drug use (IDU) history. We assessed current internet and computer access, and experience with telemedicine. We utilized logistic regression to evaluate sociodemographic associations.

**Results:** Participants’ mean (standard deviation) age was 45.9 (8.8) years, 84.0% were male, 90.1% Greek, 77.8% reported IDU within the past year, 85.2% were not linked to opioid treatment, and 50.0% were currently experiencing homelessness. Only 1.9% had experience and 46.3% had familiarity with telemedicine. Internet and computer access were reported by 66.0% and 31.5% of participants, respectively. Compared to participants with secure housing, those currently experiencing homelessness reported decreased internet (50.6% vs 81.5%, p<0.001) and computer access (11.1% vs 51.9%, p<0.001). Multivariable analyses revealed that older age (per 1-year increase: odds ratio [OR]=0.94, 95% confidence interval [CI] [0.89, 0.99], p=0.03), IDU within the past year (0.29 [0.10, 0.88], p=0.03), and homelessness (0.29, [0.13, 0.65], p=0.003) were associated with lower odds of internet access. Homelessness was associated with lower odds of computer access (0.17, [0.07, 0.41], p<0.001).

**Conclusions:** Internet and infrastructure challenges, homelessness, and digital literacy gaps should be considered to bridge the digital divide and ensure equitable digital healthcare distribution for PWUD.

Clintrials.gov registration number: NCT05794984

## Introduction

People who use drugs (PWUD) are an underserved population facing health disparities, including reduced healthcare access and discrimination within conventional medical settings.^1^ They frequently report unmet healthcare needs and predominately utilize emergency services when their health conditions deteriorate significantly.^2,3^ Healthcare inequalities for PWUD are largely driven by stigma in medical settings and other barriers, including provider mistrust, economic challenges, chaotic lifestyles, and competing priorities.^4-6^ Consequently, PWUD experience excess mortality, up to 16 times higher than the general population.^7^ Much of this excess mortality is attributed to preventable causes, such as infectious diseases, including HIV and hepatitis C virus (HCV) infections, which impose substantial burden on PWUD.^8^ Limited access to HIV and HCV treatment can greatly increase the risk of complications from these infection.^9-11^ Addressing health disparities and expanding access to medical services for PWUD are essential for improving PWUDs’ health outcomes.^12^

Digital healthcare offers a relatively innovative approach to improving healthcare access by reducing geographical and temporal barriers.^13,14^ However, the digital divide is a critical consideration for the engagement of underserved populations in digital healthcare.^15,16^ The digital divide refers to persisting healthcare disparities in digital healthcare access due largely to the influence of social determinants of health.^17^ Thus, PWUD may continue to have poor health outcomes due to reduced access to digital healthcare, mirroring the barriers to accessing healthcare in conventional health systems.^18^ While digital healthcare opens avenues for more accessible and inclusive healthcare, ensuring equitable distribution of digital healthcare is crucial when seeking to engage underserved populations.

Telehealth solutions are increasingly being investigated to provide medical and behavioral support for PWUD, particularly in the areas of HIV, HCV, substance use treatment, and mental health.^19-23^ Research indicates that telehealth interventions among PWUD have shown considerable effectiveness and high levels of patient satisfaction.^24-30^ There is a reasonable expectation that healthcare access for PWUD could be enhanced through telehealth, extending to primary care and chronic condition management, potentially addressing other preventable causes of excess morbidity and mortality. However, limited access to the internet and the requisite digital infrastructure for telehealth contribute to the digital divide for PWUD.^31^ Low digital and healthcare literacy are also important considerations for PWUD in accessing digital healthcare services.^32^ To date, there is limited research on digital healthcare accessibility among PWUD. Investigating the population’s internet and digital infrastructure access and identifying potentially influential factors are essential for tailoring digital healthcare interventions specifically for PWUD.

We aimed to explore digital healthcare accessibility among PWUD and to potentially identify factors contributing to the digital divide. Since universal healthcare systems seek to provide equitable healthcare access to all individuals, especially the underserved, and due to a lack of relevant data, we pursued this investigation in Greece.^33,34^ These findings intend to inform policy and guide the development of strategies to ensure equitable digital healthcare distribution for PWUD and other underserved populations.

## Materials and methods

Between June and July 2023, we conducted a cross-sectional study to assess digital healthcare accessibility, including internet and digital infrastructure access, as well as experience with telemedicine among PWUD in Athens, Greece. The study protocol was approved by the Institutional Review Boards of the Hellenic Scientific Society for the Study of AIDS, Sexually Transmitted and Emerging Diseases and the University at Buffalo. The study adhered to the Helsinki Declaration principles. We received and archived written informed consent from all study participants prior to their participation. We assessed, using an eligibility screener, whether participants met the inclusion criteria of being at least 18 years old, having a history of injection drug use (IDU), Greek verbal fluency, and the ability to provide informed consent. We defined PWUD as participants who had ever engaged in IDU.

### Questionnaire development

Three experienced interviewers (Z.P., E.D., V.T.) administered a structured questionnaire that included sections on sociodemographic characteristics, drug use history, utilization of conventional medical care, internet and digital infrastructure access, as well as experience with telemedicine. We based the sections on sociodemographic characteristics and history of drug use on an instrument used in a large study among PWUD in Greece.^35^ The instrument was based on the National HIV Behavioral Surveillance System for PWUD, implemented throughout the United States, and modified for relevance to Greek PWUD. We adapted the remaining questionnaire sections and questions from publications of the European Patients’ Forum and the Eurostat Model questionnaires as well as from those used in prior studies.^36-38^ Before deploying the questionnaire, we performed cognitive testing by administering the questionnaire to five study-eligible individuals. Based upon the testing results, we slightly modified the questionnaire to ensure clarity and comprehension for the study population.

### Recruitment of participants

We recruited participants through respondent-driven sampling (RDS), a chain referral methodology used to reach “hidden” or difficult-to-reach populations, in which research participation involves illegal or stigmatized behaviors.^39^ In RDS, respondents recruit their peers. Its implementation begins with a limited number of initial recruits or “seeds”.^40^ Each seed receives three coupons and is asked to recruit three additional PWUD. If the recruits are study-eligible and agree to participate, they subsequently can become recruiters, with the process continuing until the desired sample size is achieved.^41^ Recruits received a primary incentive of 10 euro for study participation followed by secondary incentives of up to 15 euro for subsequent recruitment activities.

### Questionnaire administration

The interviewers directly entered participants’ responses into a password-encrypted, secure computer database. All responses provided were participants’ self-reports. Interviewers determined participants’ housing status by asking them if they were currently living on the street, in abandoned buildings, or in shelters. Participants who responded affirmatively were grouped as participants who were currently experiencing homelessness.

### Study outcomes

The main study outcomes included digital infrastructure access, defined as current internet and computer access, as well as experience with telemedicine. We assessed the main study outcomes based on participants’ self-reported responses on the corresponding sections of the questionnaire. Secondary study outcomes included exploration of PWUDs’ perceptions about telemedicine, including perceived benefits, limitations, and willingness to engage in a telemedicine encounter.

### Statistical methods

We initially described participants’ characteristics and questionnaire responses using mean values and standard deviation (SD) or counts and percentages, as appropriate. We obtained crude and RDS-weighted (RDS-II) estimates of the main outcomes along with the corresponding 95% confidence intervals (CIs).^42^ For the 95% CIs of the weighted estimates, we used bootstrap with 1,000 replications.^43^ Since 50% of our sample was experiencing homelessness, we assessed recruitment homophily for PWUD experiencing homelessness, *i.e*., whether seeds more frequently recruited peers experiencing homelessness. Homophily values ranged from −1 to +1 (where +1 corresponds to always recruiting from one’s own group [people experiencing homelessness]) and 0 corresponds to random recruitment. We compared the responses of participants experiencing homelessness versus those with secure housing using t-tests and chi-squared tests, as appropriate. In addition, we performed univariable and multivariable analyses on the factors associated with the main study outcomes of current internet and computer access using logistic regression. Statistical calculations were performed using Stata (Stata Corp LLC. 2023. Stata Statistical Software: Release 18. College Station, TX), and p<0.05 was considered the cutoff for statistical significance.

## Results

### Sociodemographic characteristics and injection drug use

In total, 162 PWUD participated in the study (of these, 7 were RDS seeds and 155 recruits) (Table 1). One seed did not recruit any participants; out of the six remaining seeds, 3 (50.0%) were currently experiencing homelessness. PWUD currently experiencing homelessness tended to recruit other peers experiencing homelessness (homophily equal to 0.38).

**Table 1:** Sociodemographic characteristics, injecting drug use history, and participation in substance use treatment program among people who use drugs (N=162) recruited in Athens, Greece, according to housing status.

| Characteristic | Total<br>(N=162) | According to housing status |  | p |
| --- | --- | --- | --- | --- |
|  |  | Not<br>experiencing<br>homelessness<br>(N=81) | Experiencing<br>homelessness<br>(N=81) |  |
| A. Socio-demographic characteristics |  |  |  |  |
| Age (years), mean (SD) | 45.9 (8.8) | 45.6 (8.1) | 46.2 (9.6) | 0.65 |
| Sex, n (%) |  |  |  | 0.20 |
| Men | 136 (84.0) | 65 (80.3) | 71 (87.7) |  |
| Women | 25 (15.4) | 16 (19.8) | 9 (11.1) |  |
| Transgender | 1 (0.6) | 0 (0.0) | 1 (1.2) |  |
| Reside in Athens, Greece n (%) | 157 (96.9) | 77 (95.1) | 80 (98.8) | 0.08 |
| Ethnicity, n (%) |  |  |  | 0.23 |
| Greek | 146 (90.1) | 76 (93.8) | 70 (86.4) |  |
| Other | 15 (9.3) | 5 (6.2) | 10 (12.3) |  |
| Refused to answer/ don't know | 1 (0.6) | 0 (0.0) | 1 (1.2) |  |
| Employment status, n (%) |  |  |  | 0.16 |
| Employed/Student/Retired | 17 (10.5) | 12 (14.8) | 5 (6.2) |  |
| Unemployed/Unable to work | 142 (87.7) | 67 (82.7) | 75 (92.6) |  |
| Other | 3 (1.9) | 2 (2.5) | 1 (1.2) |  |
| Monthly household income<sup>1</sup>, n (%) |  |  |  | <0.001 |
| Up to 350€ | 116 (71.6) | 45 (55.6) | 71 (87.7) |  |
| 351-700€ | 19 (11.7) | 17 (21.0) | 2 (2.5) |  |
| More than 701€ | 24 (14.8) | 18 (22.2) | 6 (7.4) |  |
| Refused to answer/ don't know | 3 (1.9) | 1 (1.2) | 2 (2.5) |  |
| Marital status, n (%) |  |  |  | 0.01 |
| Single | 88 (54.3) | 38 (46.9) | 50 (61.7) |  |
| Partnered /Married | 33 (20.4) | 24 (29.6) | 9 (11.1) |  |
| Divorced/Separated/Widowed | 41 (25.3) | 19 (23.5) | 22 (27.2) |  |
| Highest education level, n (%) |  |  |  | 0.45 |
| Graduated primary school | 38 (23.5) | 17 (21.0) | 21 (25.9) |  |
| Graduated high school | 82 (50.6) | 40 (49.4) | 42 (51.9) |  |
| Graduated colleges/universities/Master/PhD | 41 (25.3) | 24 (29.6) | 17 (21.0) |  |
| Refused to answer/ don't know | 1 (0.6) | 0 (0.0) | 1 (1.2) |  |
| **B. IDU history** |  |  |  |  |
| IDU in the past 12 months, n (%) | 126 (77.8) | 64 (79.0) | 62 (76.5) | 0.71 |
| Frequency of IDU in the past 12 months, n (%) |  |  |  | 0.87 |
| At least daily | 95 (58.6) | 48 (59.3) | 47 (58.0) |  |
| Less than daily | 67 (41.4) | 33 (40.7) | 34 (42.0) |  |
| At least daily IDU in past 12 months of the following, n (%) |  |  |  |  |
| Heroin/Thai | 123 (75.9) | 63 (77.8) | 60 (74.1) | 0.95 |
| Cocaine | 59 (36.4) | 13 (16.0) | 11 (13.6) | 0.42 |
| Buprenorphine | 18 (11.1) | 8 (9.9) | 10 (12.3) | 0.60 |
| Hallucinogens (i.e., LSD) | 2 (1.2) | 2 (2.5) | 0 (0) | 0.01 |
| Sisa/Methamphetamine | 19 (11.7) | 8 (9.9) | 11 (13.6) | 0.45 |
| Speedball | 53 (32.7) | 28 (34.6) | 25 (30.9) | 0.87 |
| Sharing syringes in past 12 months, n (%) |  |  |  | 0.28 |
| No | 137 (84.6) | 71 (87.7) | 66 (81.5) |  |
| Yes | 25 (15.4) | 10 (12.4) | 15 (18.5) |  |
| **C. Participation in substance use treatment program** |  |  |  |  |
| Ever participated in a substance use treatment program, n (%) | 140 (86.4) | 71 (87.7) | 69 (85.2) | 0.65 |
| Ever participated in an OTP, n (%) | 98 (60.5) | 50 (61.7) | 48 (59.3) | 0.75 |
| Currently participate in an OTP, n (%) | 24 (14.8) | 12 (14.8) | 12 (14.8) | 0.91 |

Abbreviations: SD, standard deviation; IDU, injection drug use; LSD, Lysergic acid
diethylamide; OTP, opioid treatment program.
<sup>1</sup> For participants currently experiencing homelessness, household income was
measured as if they lived alone.

The mean (SD) age of participants was 45.9 (8.8) years, 84.0% (136/162) were male and 90.1% (146/162) were of Greek origin. In the past 12 months, 77.8% (126/162) reported IDU, and 50.0% (81/162) identified as currently experiencing homelessness. Approximately 58.6% (95/162) of participants had engaged in IDU at least daily in the past 12 months including injecting heroin (75.9%, 123/162), cocaine (36.4%, 59/162), and speedball (32.7%, 53/162). Additionally, 86.4% (140/162) of participants had previously attended a substance use treatment program and 60.5% (98/162) had participated in an opioid treatment program (OTP). However, only 14.8% (24/162) currently attended an OTP.

### Conventional medical care

PWUD most frequently obtained healthcare at public hospitals (86.4%, 140/162) (Table 2). Only 54.3% (88/162) of participants indicated that they were currently under medical care. Approximately one-third (28.4%, 46/162) of participants disclosed difficulty in accessing healthcare within the past year. Forty-three percent (70/162) of participants reported geographical barriers that required travel to another city, region, or country for healthcare. Furthermore, 56.8% (92/162) disclosed prior negative interactions with healthcare staff largely (88.0%, 81/92) attributed to previous substance use and associated stigma. Undesirable interactions included negative healthcare staff attitudes (88.0%, 81/92), refusal to administer treatment (39.1%, 36/92), and use of inappropriate language (38.0%, 35/92).

**Table 2:** Access to conventional medical care among people who use drugs (N=162) recruited in Athens, Greece, according to housing status.

| Characteristic | Total<br>(N=162) | According to housing status |  | p |
| --- | --- | --- | --- | --- |
|  |  | Not<br>experiencing<br>homelessness<br>(N=81) | Experiencing<br>homelessness<br>(N=81) |  |
| A. Conventional medical care access |  |  |  |  |
| Participants currently under care for their healthcare conditions, n (%) | 88 (54.3) | 43 (53.1) | 45 (55.6) | 0.75 |
| Participants who had visited a healthcare provider within the past 3 months, n (%) | 85 (52.5) | 37 (45.7) | 48 (59.3) | 0.08 |
| Participant rating of level of difficulty in obtaining healthcare services within the past 12 months, n (%) |  |  |  | 0.04 |
| Easy or very easy | 79 (48.8) | 39 (48.2) | 40 (49.4) |  |
| Moderate | 25 (15.4) | 16 (19.8) | 9 (11.1) |  |
| Difficult or very difficult | 46 (28.4) | 17 (21.0) | 29 (35.8) |  |
| I did not seek healthcare services within the past 12 months | 12 (7.4) | 9 (11.1) | 3 (3.7) |  |
| Where participants last accessed a healthcare provider, n (%) |  |  |  | 0.02 |
| At a public hospital | 140 (86.4) | 64 (79.0) | 76 (93.8) |  |
| At private medical practice | 12 (7.4) | 11 (13.6) | 1 (1.2) |  |
| At a health center | 5 (3.1) | 3 (3.7) | 2 (2.5) |  |
| Other | 5 (3.1) | 3 (3.7) | 2 (2.5) |  |
| B. Healthcare barriers |  |  |  |  |
| Participants who reported geographical obstacles<sup>1</sup>, n (%) | 70 (43.2) | 37 (45.7) | 33 (40.5) | 0.53 |
| Participants who reported a negative experience when accessing healthcare, n (%) | 92 (56.8) | 41 (50.6) | 51 (63.0) | 0.11 |
| Participants attributed the negative experience to, n (%) |  |  |  |  |
| Drug use history<sup>2</sup> | 81 (88.0) | 36 (87.8) | 45 (88.2) | 0.95 |
| Income/social status<sup>2</sup> | 20 (21.7) | 6 (14.6) | 14 (27.4) | 0.14 |
| Participants reported the following type of negative experience, n (%) |  |  |  |  |
| Attitude of healthcare staff<sup>2</sup> | 81 (88.0) | 37 (90.2) | 44 (86.3) | 0.56 |
| Refusal to provide me with treatment<sup>2</sup> | 36 (39.1) | 13 (31.7) | 23 (45.1) | 0.19 |
| Inappropriate language<sup>2</sup> | 35 (38.0) | 22 (53.7) | 13 (25.5) | 0.006 |
| Denial of my rights<sup>2</sup> | 16 (17.4) | 6 (14.6) | 10 (19.6) | 0.53 |

<sup>1</sup> Defined as the need to travel to another region, city, or country for healthcare.
<sup>2</sup>Calculated as proportions of the participants (N=92) who reported a negative experience when accessing healthcare.

### Digital infrastructure access

The majority (89.5%, 145/162) of study participants had previously accessed the internet, primarily through mobile phones (77.9%, 113/145) or through computers (20.0%, 29/145) (Table 3). Participants most frequently used broadband as opposed to narrowband connections. Among participants who previously accessed the internet, most (77.2%, 112/145) reported use within the past 3 months with the majority (67.9%, 76/145) reporting daily use.

**Table 3:** Technological infrastructure access among people who use drugs (N=162) recruited in Athens, Greece, according to housing status.

| Characteristic | Total<br>(N=162) | According to housing status |  | p |
| --- | --- | --- | --- | --- |
|  |  | Not<br>experiencing<br>homelessness<br>(N=81) | Experiencing<br>homelessness<br>(N=81) |  |
| Participants who had ever accessed the internet, n (%) | 145 (89.5) | 75 (92.6) | 70 (86.4) | 0.20 |
| Participants' most used device to access the internet<sup>1</sup>, n (%) |  |  |  | 0.63 |
| Mobile phone | 113 (77.9) | 58 (77.3) | 55 (78.6) |  |
| Computer | 29 (20.0) | 15 (20.0) | 14 (20.0) |  |
| Participants used the following types of internet connections<sup>1</sup>, n (%) |  |  |  |  |
| Broadband connections e.g., ADSL, VDSL, public Wi-Fi connections, mobile phone | 82 (56.5) | 56 (74.6) | 26 (37.1) | <0.001 |
| Narrowband connection e.g., via mobile phone network less than 3G | 55 (37.9) | 34 (45.3) | 21 (30.0) | 0.03 |
| Participants last used the internet<sup>1</sup>, n (%) |  |  |  | 0.03 |
| Within the last 3 months | 112 (77.2) | 64 (85.3) | 48 (68.6) |  |
| Between 3 months and one year ago | 18 (12.4) | 4 (5.3) | 14 (20.0) |  |
| More than 1 year ago | 14 (9.6) | 6 (8.0) | 8 (11.4) |  |
| Participant average use of the internet within the last 3 months<sup>2</sup>, n (%) |  |  |  | 0.66 |
| Every day or almost every day | 76 (67.9) | 45 (70.3) | 31 (64.6) |  |
| At least once a week (but not every day) | 26 (23.2) | 14 (21.9) | 12 (25.0) |  |
| Less than once a week | 9 (8.0) | 4 (6.2) | 5 (10.4) |  |
| Participants who had a current internet access, n (%) | 107 (66.0) | 66 (81.5) | 41 (50.6) | <0.001 |
| Participants who had a current computer access, n (%) | 51 (31.5) | 42 (51.9) | 9 (11.1) | <0.001 |

Abbreviations: ADSL, asymmetric digital subscriber line; VDSL, very high-speed digital
subscriber line; 3G, third generation mobile telephone.
<sup>1</sup>Calculated as proportions of the participants (N=145) who reported ever access to the internet.
<sup>2</sup>Calculated as proportions of the participants (N=112) who reported last use of the internet within the past 3 months.

Among all participants, approximately two-thirds (66.0%, 107/162, 95% CI [58.2 – 73.3]) reported current internet access (RDS-weighted prevalence: 68.7%, 95% CI [53.7 – 81.3]). Compared to participants with secure housing, we observed significantly lower current internet access among those currently experiencing homelessness (81.5%, 66/81 vs. 50.6%, 41/81, p<0.001). In multivariable analysis, we observed that current homelessness (0.29, 95% CI: [0.13, 0.65], p=0.003), increasing age (per 1-year increase: 0.94, 95% CI: [0.89, 0.99], p=0.03) and IDU within the past 12 months (0.29, 95% CI: [0.10, 0.88], p=0.03) were associated with decreased odds of current internet access (Table 4).

**Table 4.** Logistic regression results to identify factors associated with current internet and computer access among people who use drugs (N=162) recruited in Athens, Greece.

|  | Odds Ratio | 95% CI | p |
| --- | --- | --- | --- |
| **A. Current internet access** |  |  |  |
| Age | 0.94 | [0.89, 0.99] | 0.03 |
| In the past 12 months, did you use intravenous drugs other than those prescribed to you for medical reasons? |  |  |  |
| No | 1.00 |  |  |
| Yes | 0.29 | [0.10, 0.88] | 0.03 |
| Are you currently experiencing homelessness?<sup>1</sup> |  |  |  |
| No | 1.00 |  |  |
| Yes | 0.29 | [0.13, 0.65] | 0.003 |
| How many years have you attended school? | 1.13 | [0.96, 1.33] | 0.15 |
| What is your ethnicity? |  |  |  |
| Greek | 1.00 |  |  |
| Non Greek | 1.23 | [0.31, 4.81] | 0.77 |
| What best describes your employment status? |  |  |  |
| Employed/Student/Retired | 1.00 |  |  |
| Unemployed/Unable to work | 0.63 | [0.17, 2.33] | 0.49 |
| Marital status |  |  |  |
| Single | 1.00 |  |  |
| Relationship/Married | 1.38 | [0.50, 3.86] | 0.54 |
| Divorced/Separated/Widowed | 1.59 | [0.60, 4.21] | 0.35 |
| **B. Current computer access** |  |  |  |
| Age | 0.94 | [0.88, 1.00] | 0.05 |
| In the past 12 months, did you use intravenous drugs other than those prescribed to you for medical reasons? |  |  |  |
| No | 1.00 |  |  |
| Yes | 1.90 | [0.60, 6.07] | 0.28 |
| Are you currently experiencing homelessness?<sup>1</sup> |  |  |  |
| No | 1.00 |  |  |
| Yes | 0.17 | [0.07, 0.41] | <0.001 |
| How many years have you attended school? | 1.10 | [0.90, 1.35] | 0.36 |
| What is your ethnicity? |  |  |  |
| Greek | 1.00 |  |  |
| Non Greek | 0.42 | [0.08, 2.28] | 0.32 |
| What best describes your employment status? |  |  |  |
| Employed/Student/Retired | 1.00 |  |  |
| Unemployed/Unable to work | 0.25 | [0.07, 0.98] | 0.05 |
| Marital status |  |  |  |
| Single | 1.00 |  |  |
| Relationship/Married | 1.79 | [0.68,<br>4.76] | 0.24 |
| Divorced/Separated/Widowed | 0.77 | [0.25,<br>2.33] | 0.64 |

Abbreviations: CI, Confidence interval
<sup>1</sup>Currently experiencing homelessness defined as currently living on the street, in abandoned buildings, or in shelters.

Overall, 31.5% (51/162) of participants reported current computer access. Compared to participants with secure housing, we observed significantly lower current computer access among those experiencing homelessness (51.9%, 42/81 vs. 11.1%, 9/81, p<0.001). In multivariate analysis, we observed that current homelessness (0.17 [0.07, 0.41], p<0.001) was associated with decreased odds of current computer access (Table 4).

### Experience with telemedicine

Very few participants had ever used (1.9%, 3/162), and less than half (46.3%, 75/162) had familiarity with telemedicine (Supplemental file 1). Initially, most (71.0%, 115/162) participants indicated their preference to participate in telemedicine encounters in their homes. When provided the choice of participating in a telemedicine encounter in an OTP, most (136/162, 84.0%) participants endorsed receiving care through telemedicine in that setting.

Participants perceived telemedicine to be of high value due to its time-saving nature by eliminating the need to travel to an appointment (87.0%, 141/162), the convenience of being able to participate from anywhere (84.6%, 137/162), reduced provider wait times (84.0%, 136/162), and reduced infection exposure risk by avoiding in-person visits (79.0%, 128/162) (Supplemental files 2 and 3). Most participants (76.5%, 124/162) perceived the inability to perform a physical examination as telemedicine’s leading limitation. Other perceived limitations of telemedicine included the lack of direct personal contact (69.8%, 113/162), the need to use digital infrastructure for telemedicine participation (64.2%, 104/162), and potential technical issues (61.1%, 99/162). Difficulty trusting the doctor (39.5%, 64/162) and substandard patient-doctor relationships (35.8%, 58/162) were the least frequently perceived telemedicine limitations (Supplemental files 4 and 5).

## Discussion

We pursued this investigation to evaluate digital healthcare accessibility among PWUD in Athens, Greece. We found that the vast majority of study participants lacked experience with telemedicine, consistent with results obtained from most European countries.^44^ When we investigated digital infrastructure accessibility, we found that two-thirds of participants reported currently connecting to the internet, while one-third reported current computer access. In multivariate analysis, we identified that current homelessness, older age, and IDU within the past 12 months were associated with significantly reduced internet and/or computer access among PWUD. These findings highlight important considerations for expansion of digital health among underserved populations. Figure 1 describes several considerations for utilizing digital approaches to distribute healthcare equitably to underserved populations.

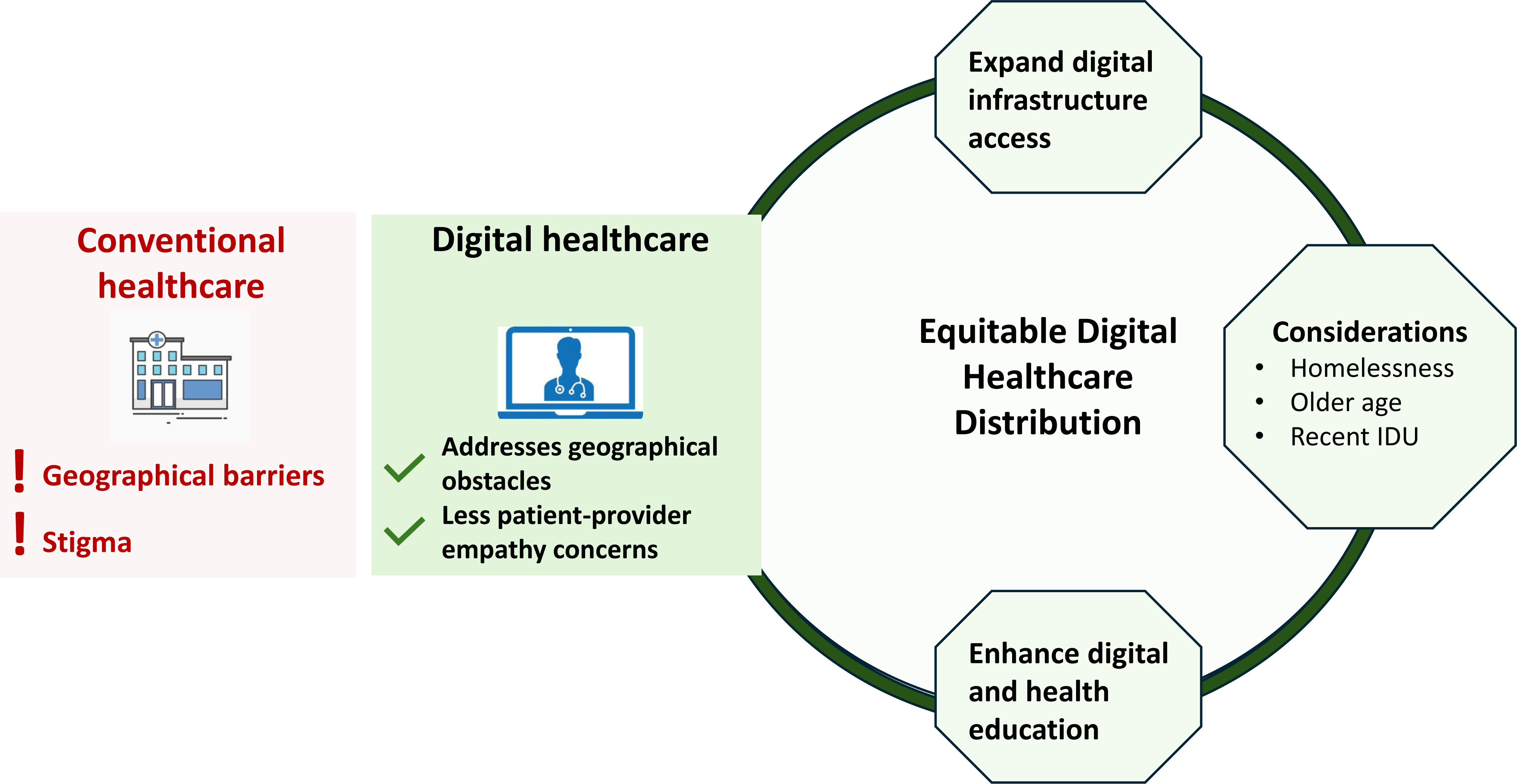
Figure legend: Promotion of Equitable Digital Healthcare Distribution. People who use drugs (PWUD) are a medically underserved population who reported encountering geographical obstacles and stigma in accessing healthcare in conventional settings. Digital healthcare simultaneously overcomes geographical obstacles accompanied by few empathy concerns. Therefore, the equitable distribution of digital healthcare services to PWUD is critical to avoid worsening future healthcare access. In our investigation we found that homelessness, increasing age, and injection drug use within the past 12 months were associated with reduced internet and digital infrastructure access. We also identified gaps in digital literacy. To promote equitable distribution of digital healthcare, thereby ensuring health equity, requires expansion of internet and digital infrastructure access for underserved populations and addressing digital literacy gap. Abbreviations: IDU, injection drug use.

One third of participants reported difficulty accessing healthcare within the past year and only approximately half indicated that they were under medical care at the time of the questionnaire administration. Geographic barriers and stigma were frequently cited obstacles to healthcare access in conventional healthcare venues, consistent with prior studies.^45-47^ Study participants were more concerned with digital infrastructural challenges rather than with the expression of empathy through telemedicine. These perceptions align with findings from both a staff-facilitated telemedicine model for HCV care integrated into OTPs and a peer-facilitated telehealth model for HIV care conducted in syringe services programs. In both of these interventions participants highly valued empathy, trust, and telehealth’s destigmatizing approach.^27,48^ Considering that stigma is a significant barrier to healthcare access for PWUD, engendering trust and empathy are key for effective healthcare delivery, including through digital approaches.^5,27,49^ With a separate group of PWUD in Athens Greece, we conducted focus groups for an indepth understanding of barriers and facilitators of their digital health use. Focus group participants suggested that an initial in-person appointment, eye contact during telehealth encounters, partnerships with PWUD-supportive community organizations, and patient education could enhance trust in telehealth.^50^ Our current investigation also identified the need to enhance patient education and digital literacy, as less than half of the participants had familiarity with telemedicine.

Internet access and digital infrastructure are absolute requirements for digital health participation, yet limited data exist on digital infrastructure access among PWUD. A previous survey of 204 PWUD in Greece revealed that over 90.0% had internet access.^51^ Participants in that study were recruited from substance use treatment programs, which may not represent the real-world experiences of PWUD outside those supportive environments. In contrast, only 14.8% (24/162) of participants in our study were enrolled in an OTP. Since IDU is a potential transmission route for HCV and HIV, tailored telehealth interventions should be directed toward PWUD.^52^ Our study sample, of whom the majority (77.8%, 126/162) disclosed IDU within the past 12 months, is representative of the PWUD population that should be prioritized in subsequent telehealth interventions.

Our study sample included a substantial proportion currently experiencing homelessness (50.0%), a difficult population to enroll in research.^53^ The homelessness percentage was higher than in other studies among a similar Athenian population, where rates of homelessness ranged from 23.1% to 25.6%.^35,53^ The inclusion of PWUD experiencing homelessness as seeds (*i.e.,* three of six actively recruiting study seeds), the moderate homophily among participants experiencing homelessness and the small sample size account for the over-representation of this population subgroup. Nonetheless, the recruitment of many participants experiencing homelessness enabled us to investigate their digital healthcare accessibility and insights into telemedicine for the first time. As expected, we found that homelessness is associated with reduced access to both conventional and digital healthcare.

When designing interventions to ensure equitable distribution of digital healthcare to PWUD, approaches inclusive of those experiencing homelessness are necessary.^30,54^ Our study participants indicated that they predominately used mobile phones to access the internet, consistent with another study in which over 75.0% of participants used mobile phones for this purpose.^55^ In that study, PWUD participants were provided mobile phones through a government-supported program for low-income individuals. These findings highlight the government’s central role in supporting digital healthcare access among underserved populations. The frequent turnover of government-issued devices due to theft or resale by PWUD is an important consideration when assessing their worthiness. Fleeting device ownership may limit their suitability for digital healthcare access by PWUD. Additionally, restricting the phone’s functionality to solely that required for telehealth encounters may discourage its use. Other sociodemographic factors, such as increasing age and unemployment, could further affect PWUDs’ ability to leverage the benefits of digital healthcare.^56^ A potential solution to bridging the digital divide for healthcare delivery to underserved populations could be the expansion of the facilitated telemedicine model, where digital healthcare is integrated into convenient locations for PWUD, such as OTPs, and is supported by case managers.^25,57^ Most study participants expressed a willingness to receive care through telemedicine in an OTP.

Of the study limitations, the most important is that our sample consists of PWUD recruited from downtown Athens, which may not be representative of PWUD populations in other geographical areas. Participants’ responses, including the prevalence of HCV and HIV, were self-reported, introducing potential ascertainment bias.^58^ Furthermore, individuals experiencing homelessness are more susceptible to acquiring HIV and HCV, which may have resulted in the high self-reported prevalences observed in our study population.^59^

## Conclusions

PWUD who are currently experiencing homelessness accessed the internet and computers at significantly lower rates compared to those with stable housing. In addition to homelessness, older age, and IDU within the past 12 months were associated with reduced access to the necessary infrastructure for telehealth participation. These factors exacerbate the digital divide among PWUD and should be considered, along with approaches to bridge digital literacy gaps, when designing digital healthcare interventions for PWUD. Further studies are needed to investigate implementation approaches for digital healthcare delivery to underserved populations. Expanding internet and digital infrastructure access is essential to promote health equity in the distribution of digital healthcare to PWUD.

## Data Availability

The study data are available from the corresponding author upon reasonable request.

## Acknowledgments

We thank the study participants for their involvement with the study and the staff of Prometheus for enabling us to utilize their site.

## Authors’ contribution statement

Z.P.: writing – original draft, project administration (lead); investigation (lead); supervision (lead); visualization (lead). S.R.: formal analysis (equal); software (lead); writing – review and editing (equal). E.D.: resources (equal); writing – review and editing (supporting). V.T.: resources (equal); writing – review and editing (supporting). G.K.: resources (equal); writing – review and editing (supporting). A.D.: writing – review and editing (supporting). V.S.: formal analysis (equal); software (supporting); writing – review and editing (supporting). A.H.: investigation (lead); methodology (lead); supervision (lead); visualization (lead); writing – review and editing (supporting). A.H.T.: investigation (lead); methodology (lead); supervision (lead); visualization (lead); funding acquisition; writing – review and editing (equal). All authors approved the final version of the manuscript.

## Data Sharing

The study data are available from the corresponding author upon reasonable request.

---

**Supplemental file 1.** Telemedicine experience and perceptions among people who use drugs (N=162) recruited in Athens, Greece, according to housing status.

| Characteristic | Total<br>(N=162) | According to housing status |  | p |
| --- | --- | --- | --- | --- |
|  |  | Not<br>experiencing<br>homelessness<br>(N=81) | Experiencing<br>homelessness<br>(N=81) |  |
| Participants who had ever heard about telemedicine, n (%) | 75 (46.3) | 45 (55.6) | 30 (37.0) | 0.02 |
| Participants who had ever used telemedicine, n (%) | 3 (1.9) | 2 (2.5) | 1 (1.2) | 1.00 |
| Participants who find telemedicine concept interesting, n (%) | 98 (60.5) | 49 (60.5) | 49 (60.5) | 1.00 |
| Participant perceptions about telemedicine versus conventional medical care, n (%) |  |  |  | 0.65 |
| Conventional medical care is superior | 89 (54.9) | 45 (55.6) | 44 (54.3) |  |
| Telemedicine is superior | 29 (17.9) | 16 (19.8) | 13 (16.0) |  |
| It has no difference | 32 (19.8) | 16 (19.8) | 16 (19.8) |  |
| Refused/don't know | 12 (7.4) | 4 (4.9) | 8 (9.9) |  |
| Participant preferred location to receive telemedicine, n (%) |  |  |  | 0.64 |
| OTP | 36 (22.2) | 19 (23.5) | 17 (21.0) |  |
| Home | 115 (71.0) | 56 (69.1) | 59 (72.8) |  |
| Internet café | 2 (1.2) | 2 (2.5) | 0 (0.0) |  |
| Participant willingness to receive telemedicine care in an OTP, n (%) | 136 (84.0) | 64 (79.0) | 72 (88.9) | 0.09 |

Abbreviation: OTP, opioid treatment program.

**Supplemental file 2.** Potential benefits of telemedicine, as identified by people who use drugs (N=162) recruited in Athens, Greece, and according to housing status. Participants were allowed to choose more than one response.

| Characteristic | Total<br>(N=162) | According to housing status |  | p |
| --- | --- | --- | --- | --- |
|  |  | Not<br>experiencing<br>homelessness<br>(N=81) | Experiencing<br>homelessness<br>(N=81) |  |
| Convenience (e.g., can do it from home), n (%) | 137 (84.6) | 73 (90.1) | 64 (79.0) | 0.05 |
| Time-saving (not having to travel to an appointment), n (%) | 141 (87.0) | 73 (90.1) | 68 (84.0) | 0.24 |
| Reduce travel expenses, n (%) | 123 (75.9) | 67 (82.7) | 56 (69.1) | 0.04 |
| Not waiting as long to be seen for an ailment, n (%) | 136 (84.0) | 68 (84.0) | 68 (84.0) | 1.00 |
| Not paying as much, n (%) | 84 (51.9) | 44 (54.3) | 40 (49.4) | 0.53 |
| Higher level of flexibility, n (%) | 109 (67.3) | 56 (69.1) | 53 (65.4) | 0.62 |
| Reduction of risk for infections by avoiding in-person visits, n (%) | 128 (79.0) | 68 (84.0) | 60 (74.1) | 0.12 |
| More efficient in interactions with a healthcare provider, n (%) | 82 (50.6) | 44 (54.3) | 38 (46.9) | 0.35 |
| Treated with respect (non-judgmental), n (%) | 95 (58.6) | 51 (63.0) | 44 (54.3) | 0.26 |

**Supplemental file 3.**
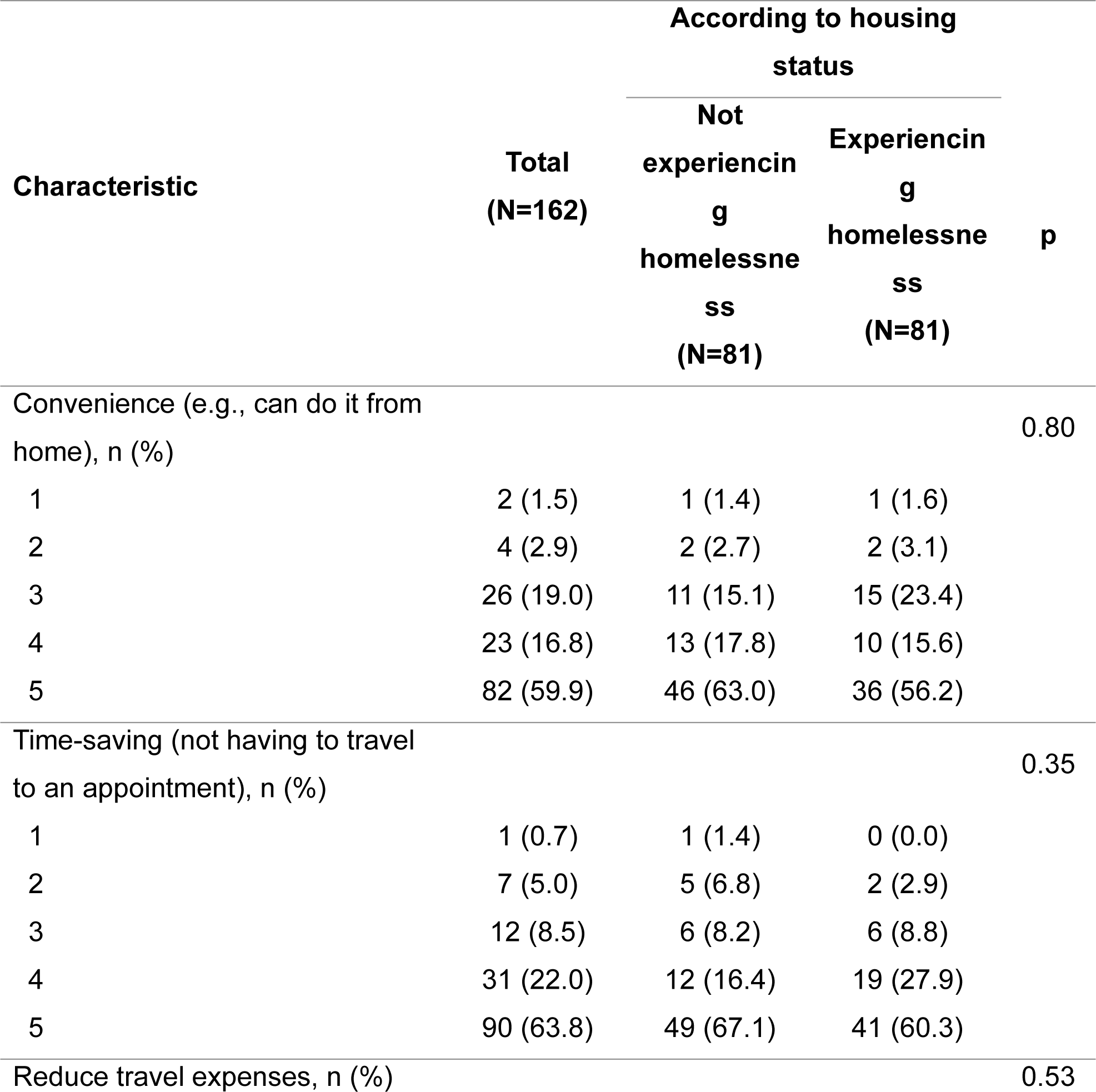
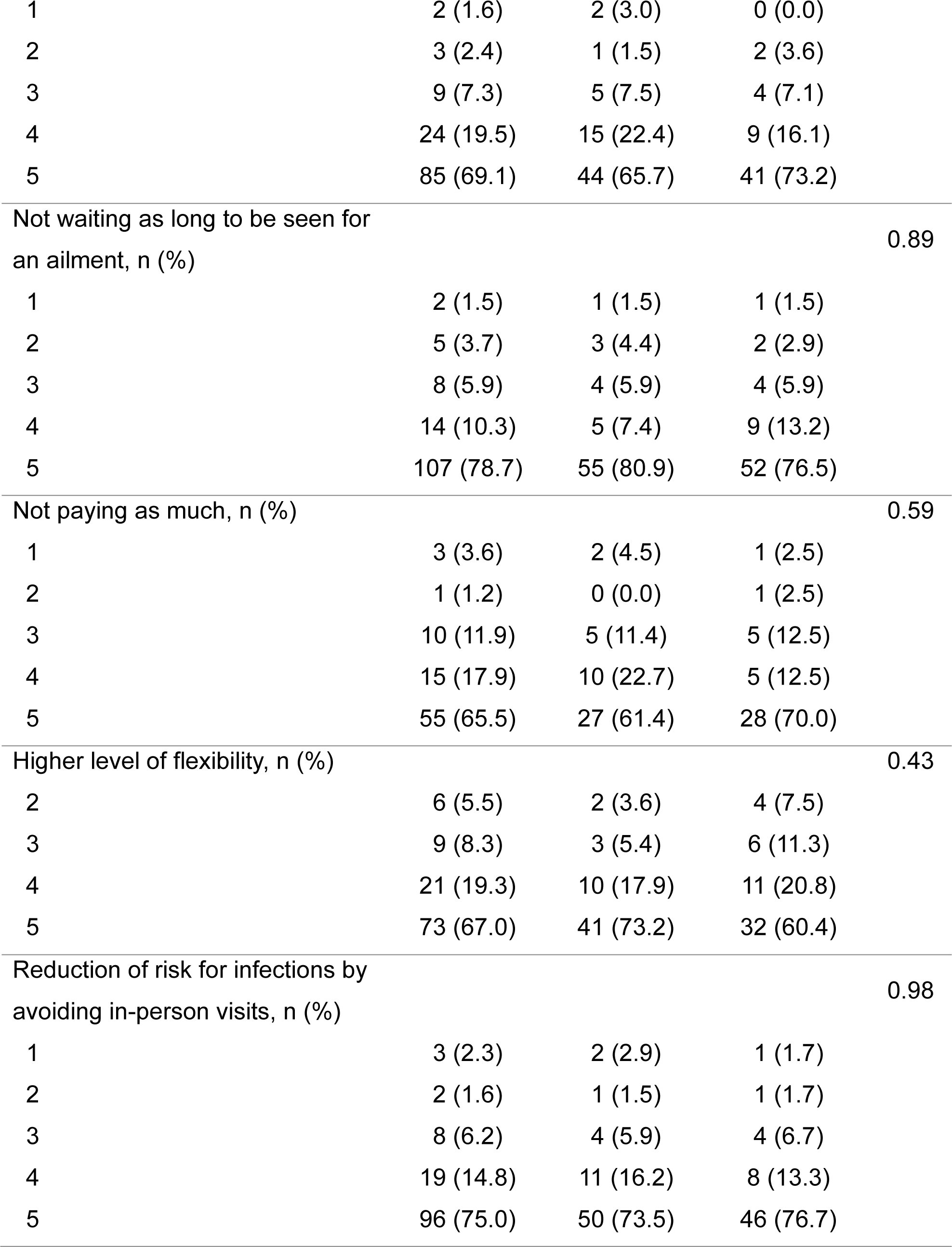
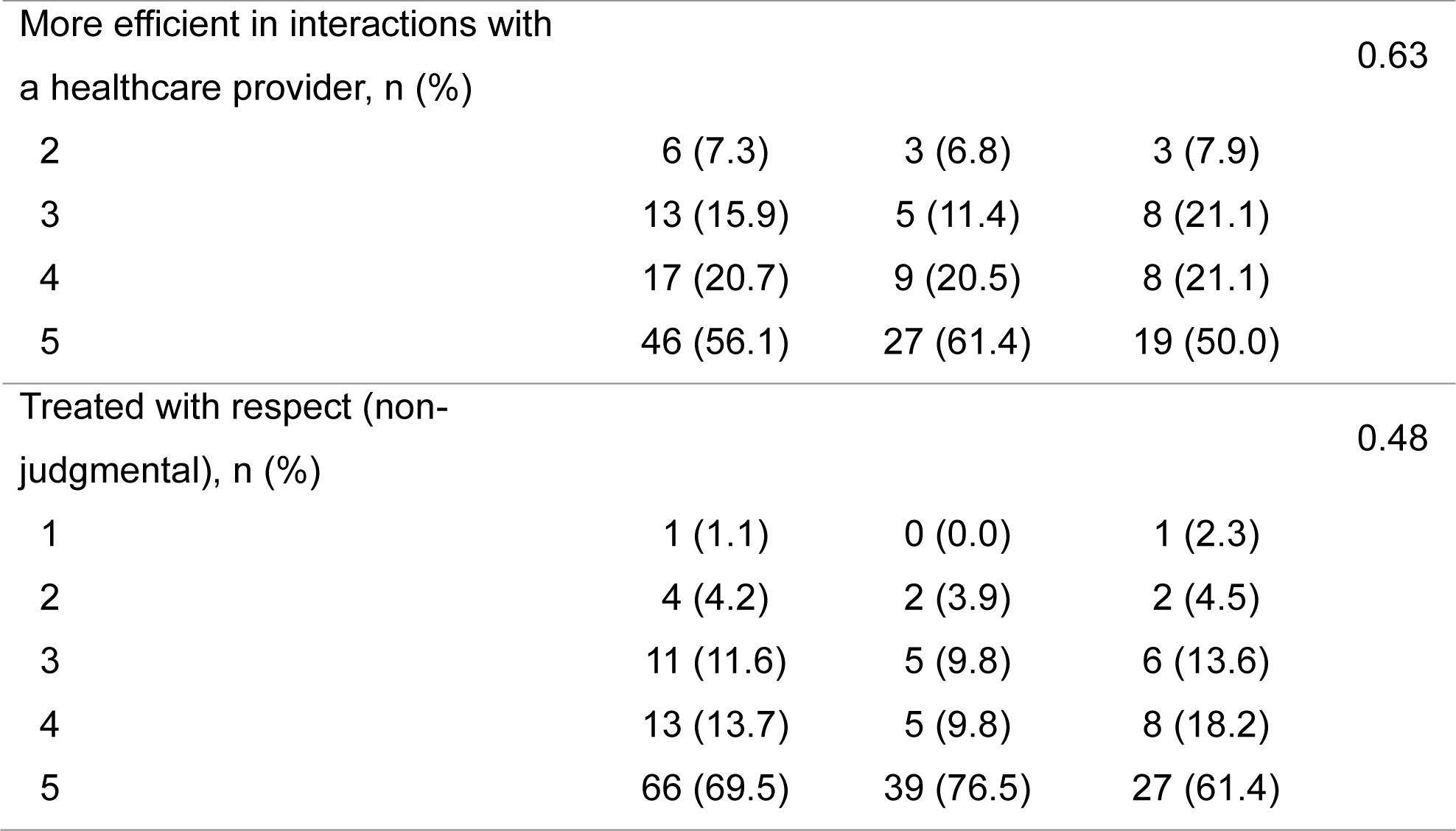
Rating on a scale from 1-5 (where 1=minimum and 5=maximum) of every potential benefit of telemedicine chosen in the previous question, according to the importance to them, among people who use drugs (N=162) recruited in Athens, Greece, and according to housing status.

**Additional file 4.** Potential limitations of telemedicine, as identified by people who use drugs (N=162) recruited in Athens, Greece, and according to housing status. Participants were allowed to choose more than one response.

| Characteristic | Total<br>(N=162) | According to housing status |  | p |
| --- | --- | --- | --- | --- |
|  |  | Not<br>experiencing<br>homelessness<br>(N=81) | Experiencing<br>homelessness<br>(N=81) |  |
| Privacy issues, n (%) | 94 (58.0) | 45 (55.6) | 49 (60.5) | 0.52 |
| Security threats, n (%) | 82 (50.6) | 41 (50.6) | 41 (50.6) | 1.00 |
| Technical problems, n (%) | 99 (61.1) | 55 (67.9) | 44 (54.3) | 0.08 |
| Requires technology devices, n (%) | 104 (64.2) | 56 (69.1) | 48 (59.3) | 0.19 |
| No direct personal contact, n (%) | 113 (69.8) | 59 (72.8) | 54 (66.7) | 0.39 |
| No physical examination possible, n (%) | 124 (76.5) | 60 (74.1) | 64 (79.0) | 0.46 |
| Difficulty trusting the doctor, n (%) | 64 (39.5) | 37 (45.7) | 27 (33.3) | 0.11 |
| Substandard patient-doctor relationship, n (%) | 58 (35.8) | 30 (37.0) | 28 (34.6) | 0.74 |

**Additional file 5.**
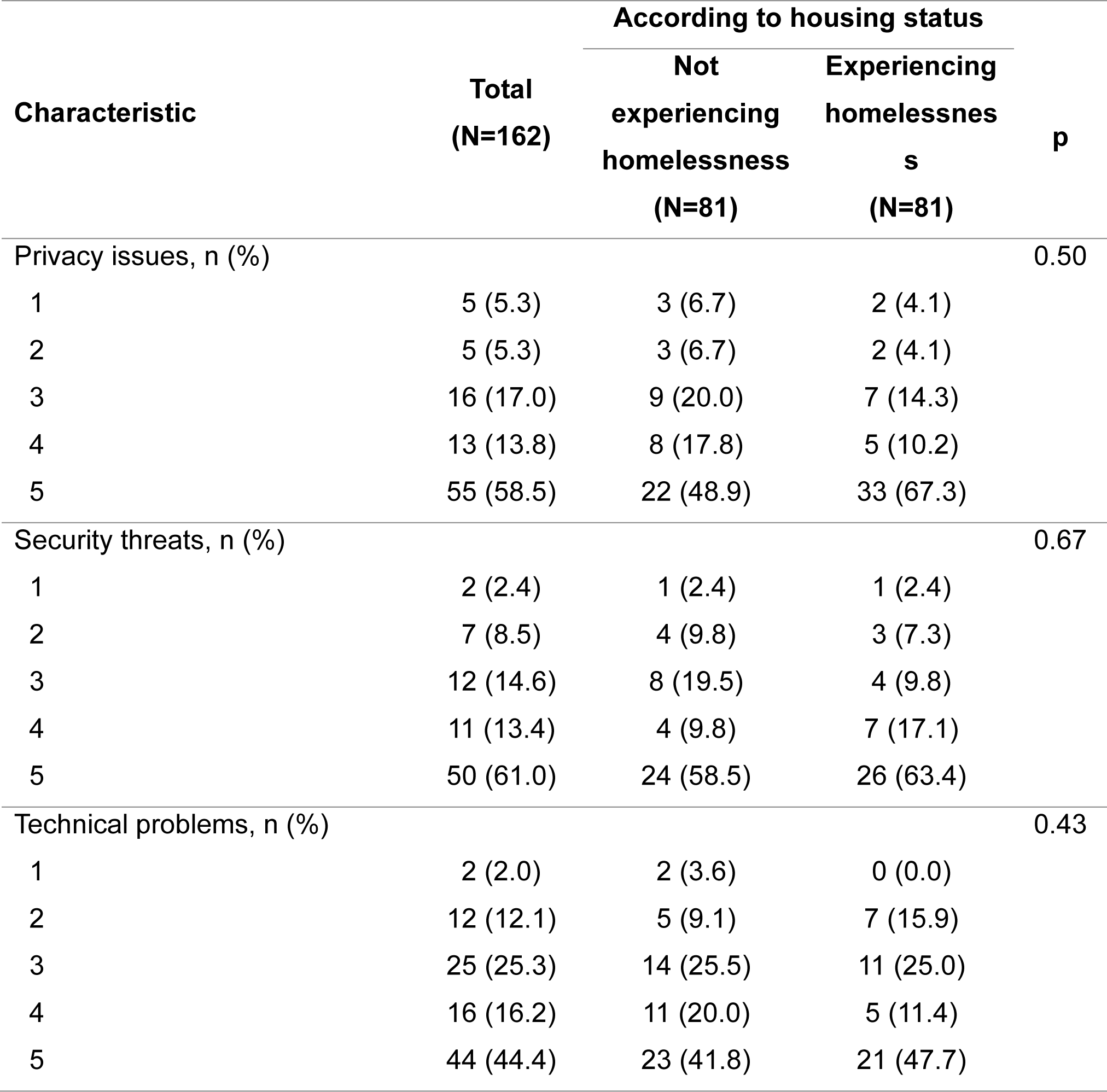
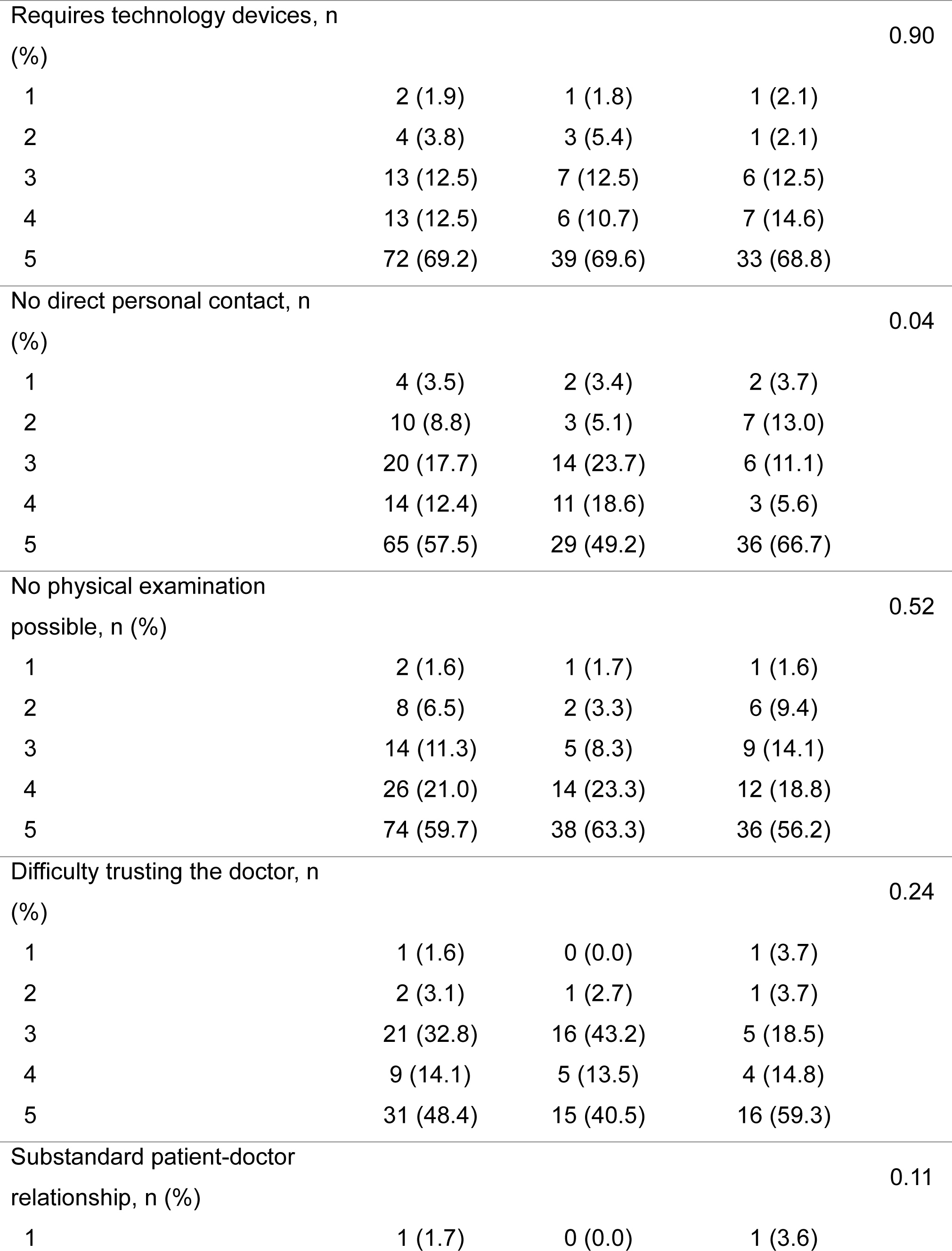
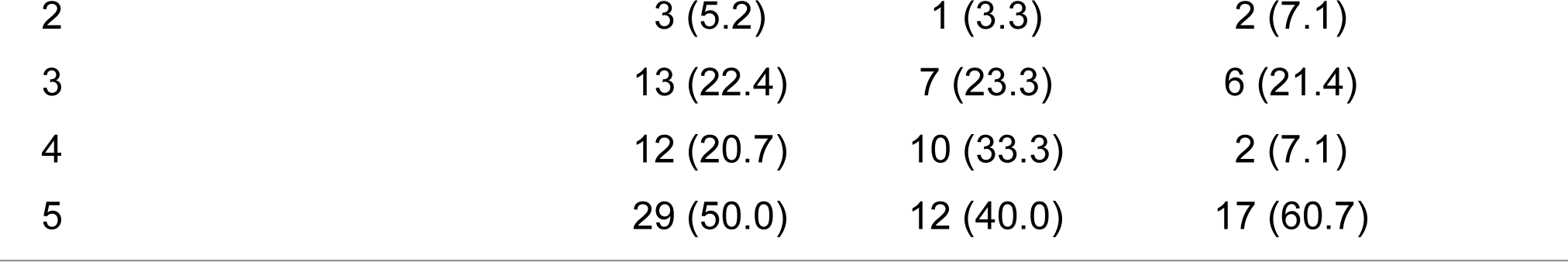
Rating on a scale from 1-5 (where 1=minimum and 5=maximum) of every potential limitation of telemedicine chosen in the previous question, according to the importance to them, among people who use drugs (N=162) recruited in Athens, Greece, and according to housing status.

| Characteristic | Total<br>(N=162) | According to housing status |  | p |
| --- | --- | --- | --- | --- |
|  |  | Not<br>experiencing<br>homelessness<br>(N=81) | Experiencing<br>homelessness<br>(N=81) |  |
| Privacy issues, n (%) |  |  |  | 0.50 |
| 1 | 5 (5.3) | 3 (6.7) | 2 (4.1) |  |
| 2 | 5 (5.3) | 3 (6.7) | 2 (4.1) |  |
| 3 | 16 (17.0) | 9 (20.0) | 7 (14.3) |  |
| 4 | 13 (13.8) | 8 (17.8) | 5 (10.2) |  |
| 5 | 55 (58.5) | 22 (48.9) | 33 (67.3) |  |
| Security threats, n (%) |  |  |  | 0.67 |
| 1 | 2 (2.4) | 1 (2.4) | 1 (2.4) |  |
| 2 | 7 (8.5) | 4 (9.8) | 3 (7.3) |  |
| 3 | 12 (14.6) | 8 (19.5) | 4 (9.8) |  |
| 4 | 11 (13.4) | 4 (9.8) | 7 (17.1) |  |
| 5 | 50 (61.0) | 24 (58.5) | 26 (63.4) |  |
| Technical problems, n (%) |  |  |  | 0.43 |
| 1 | 2 (2.0) | 2 (3.6) | 0 (0.0) |  |
| 2 | 12 (12.1) | 5 (9.1) | 7 (15.9) |  |
| 3 | 25 (25.3) | 14 (25.5) | 11 (25.0) |  |
| 4 | 16 (16.2) | 11 (20.0) | 5 (11.4) |  |
| 5 | 44 (44.4) | 23 (41.8) | 21 (47.7) |  |
| Requires technology devices, n (%) |  |  |  | 0.90 |
| 1 | 2 (1.9) | 1 (1.8) | 1 (2.1) |  |
| 2 | 4 (3.8) | 3 (5.4) | 1 (2.1) |  |
| 3 | 13 (12.5) | 7 (12.5) | 6 (12.5) |  |
| 4 | 13 (12.5) | 6 (10.7) | 7 (14.6) |  |
| 5 | 72 (69.2) | 39 (69.6) | 33 (68.8) |  |
| No direct personal contact, n (%) |  |  |  | 0.04 |
| 1 | 4 (3.5) | 2 (3.4) | 2 (3.7) |  |
| 2 | 10 (8.8) | 3 (5.1) | 7 (13.0) |  |
| 3 | 20 (17.7) | 14 (23.7) | 6 (11.1) |  |
| 4 | 14 (12.4) | 11 (18.6) | 3 (5.6) |  |
| 5 | 65 (57.5) | 29 (49.2) | 36 (66.7) |  |
| No physical examination possible, n (%) |  |  |  | 0.52 |
| 1 | 2 (1.6) | 1 (1.7) | 1 (1.6) |  |
| 2 | 8 (6.5) | 2 (3.3) | 6 (9.4) |  |
| 3 | 14 (11.3) | 5 (8.3) | 9 (14.1) |  |
| 4 | 26 (21.0) | 14 (23.3) | 12 (18.8) |  |
| 5 | 74 (59.7) | 38 (63.3) | 36 (56.2) |  |
| Difficulty trusting the doctor, n (%) |  |  |  | 0.24 |
| 1 | 1 (1.6) | 0 (0.0) | 1 (3.7) |  |
| 2 | 2 (3.1) | 1 (2.7) | 1 (3.7) |  |
| 3 | 21 (32.8) | 16 (43.2) | 5 (18.5) |  |
| 4 | 9 (14.1) | 5 (13.5) | 4 (14.8) |  |
| 5 | 31 (48.4) | 15 (40.5) | 16 (59.3) |  |
| Substandard patient-doctor relationship, n (%) |  |  |  | 0.11 |
| 1 | 1 (1.7) | 0 (0.0) | 1 (3.6) |  |

|  |  |  |  |
| --- | --- | --- | --- |
| 2 | 3 (5.2) | 1 (3.3) | 2 (7.1) |
| 3 | 13 (22.4) | 7 (23.3) | 6 (21.4) |
| 4 | 12 (20.7) | 10 (33.3) | 2 (7.1) |
| 5 | 29 (50.0) | 12 (40.0) | 17 (60.7) |

